# Improved symptoms following bumetanide treatment in children aged 3 to 6 years with autism spectrum disorder: a randomized, double-blind, placebo-controlled trial

**DOI:** 10.1101/2020.09.18.20197640

**Authors:** Yuan Dai, Lingli Zhang, Juehua Yu, Xin Zhou, Hua He, Yiting Ji, Kai Wang, Xiujuan Du, Xin Liu, Yun Tang, Shining Deng, Christelle Langley, Wei-Guang Li, Jun Zhang, Jianfeng Feng, Barbara J Sahakian, Qiang Luo, Fei Li

## Abstract

With the drug therapy for the core symptoms of autism spectrum disorder (ASD) currently limited, here we reported a randomised, double-blind, placebo-controlled trial to investigate the efficacy, safety, and potential neural mechanism of bumetanide in children with ASD aged 3 to 6 years old. There were 120 children entered into the study and randomly assigned to either 0.5mg bumetanide or placebo. In the final sample, 119 received at least one dose of bumetanide (59 children) or placebo (60 children). The primary outcome was the score reduction of Childhood Autism Rating Scale (CARS) and the secondary outcomes were the score of Clinical Global Impressions Scale (CGI) -Global Improvement (CGI-I) at 3 months and the change from baseline to 3-month in Autism Diagnostic Observation Schedule (ADOS). Magnetic resonance spectroscopy (MRS) was used to measure γ-aminobutyric acid (GABA) and glutamate neurotransmitter concentrations in the insular cortex (IC) before and after the treatment. As compared with the placebo, bumetanide treatment was significantly better in reducing severity. No patient withdrew from the trial due to adverse events. The superiority of bumetanide to placebo in reducing insular GABA, measured using MRS, was demonstrated. The clinical improvement was associated with the decrease in insular GABA in the bumetanide group. In conclusion, this trial in a large group of young children with predominantly moderate and severe ASD demonstrated that bumetanide is safe and effective in improving the core symptoms of ASD. However, the clinical significance remains uncertain and future multi-center clinical trials are required to replicate these findings and confirm the clinical significance using a variety of outcome measures.

## Introduction

Autism spectrum disorder (ASD) is an early-onset neurodevelopmental disorder and often leads to a life-time disability, and currently has no cure and no U.S. Food and Drug Administration (FDA)-approved medication to effectively treat its core symptoms including social communication and social interaction as well as restricted, repetitive patterns of behaviour ^[1-3]^. Behavioural interventions in early childhood are most effective and recommended ^[4, 5]^. However, behaviour therapy services are not widely available for early childhood intervention in many countries ^[6, 7]^.

Bumetanide, an FDA-approved potent loop diuretic, has been proposed as a promising drug candidate for ASD treatment. In previous studies the use of bumetanide was shown to improve the symptoms including social communication, interaction, and restricted interest and it was also shown to attenuate the severity of the disorder in ASD patients, as measured by Childhood Autism Rating Scale (CARS,) with no major adverse events ^[8-11]^. The first randomised controlled trial in 60 cases aged 3 to 11 years, of which the average age was 6.8 years, showed improvements in CARS and Clinical Global Impressions Scale (CGI)-Global Improvement scale (CGI-I) with a main side effect mild hypokalemia ^[12]^. Another randomised controlled unblinded trial on 60 cases aged 2.5 to 6.5 years found that the add-on bumetanide together with the Applied Behaviour Analysis (ABA) was superior to ABA alone as measured by both CGI-I and Autism Behaviour Checklist (ABC) total score. In that study, no significant side effects were reported^[13]^. A randomised, four-arm, dose-response trial to optimize bumetanide dosage on 88 cases aged 2 to 18 years showed improvements in CARS total score and global Social Responsiveness Scales (SRS) score, as well as the subcategories social communication and restricted interests and repetitive behaviour in SRS, however some adverse events were observed, including hypokalemia, increased urine elimination, loss of appetite, dehydration and asthenia^[14]^. Limitations of these trials included relatively small sample sizes and the patient heterogeneity in terms of a large age range from infants to adolescents. Therefore, the safety and efficacy of bumetanide in young children remain uncertain. In addition, it is important to understand the neural mechanisms of symptom reduction by bumetanide in the human autistic brain. Towards filling this gap, our most recent open-label trial on 83 children with ASD aged 3 to 6 years showed improvements in both CARS and CGI and provided the neuroimaging evidence suggesting that bumetanide reduces γ-aminobutyric acid (GABA) in the human autistic brain, especially in the insular cortex (IC)^[15]^. The IC is known as a brain region involved in the integration of sensory, emotional, and autonomic information in order to allocate cognitive resources and guide behaviour^[16]^. These functions were variably impaired in ASD rodent models and in individuals with ASD^[17]^. Our previous findings suggested that bumetanide decreases GABA in the IC, and thereby ameliorates ASD symptoms in children^[15]^.

These recent findings by our group encouraged us to conduct a randomised, double-blind, placebo-controlled trial of bumetanide, in which we investigated the safety and efficacy of bumetanide treatment for 3 months in children aged 3 to 6 years with ASD. In addition, we used magnetic resonance spectroscopy (MRS) to measure the neurotransmitter concentrations of GABA and glutamate in the IC obtained before and after treatment. This would allow us to determine whether the therapeutic effect of bumetanide on autistic symptoms was associated with changes in GABA.

## Materials and methods

Detailed methods are provided in the Appendix-supplementary methods.

### Trial design and participants

The study used a randomised, double-blind, placebo-controlled, parallel-group design. We recruited eligible children from outpatient clinics at the Xinhua Hospital affiliated to Shanghai Jiao Tong University School of Medicine. Participants were considered eligible if they were diagnosed with ASD according to the Diagnostic and Statistical Manual of Mental Disorders, Fifth Edition (DSM-5); aged between 3 to 6 years; confirmed diagnosis with the Autism Diagnostic Interview–Revised (ADIR) and/or Autism Diagnostic Observation Schedule (ADOS), a CARS total score of no less than 30 and had no access to any behavioural intervention. We excluded patients if they had liver or kidney dysfunction; a history of allergy to sulfa drugs; abnormal electrocardiogram; genetic or chromosomal abnormalities (exome-sequencing and/or microarray assays); were diagnosed with neurological disease (e.g. epilepsy, Rett syndrome), or psychiatric disorder (e.g. very early-onset schizophrenia) other than ASD; severe hearing or visual impairment; were currently using melatonin for the treatment of sleep disorders or cessation of such treatment for less than 3 weeks. Additional exclusion criteria for neuroimaging were any contraindications of magnetic resonance imaging (MRI) scanning and any previous reports of traumatic brain injury.

Participants were randomly assigned in a 1:1 ratio to receive 0.5mg oral bumetanide or placebo twice daily for 3 months by use of a block randomisation scheme. Bumetanide and placebo tablets were identical in appearance, smell and taste. Patients and their caregivers, investigators, experienced psychiatrists, and data analysts remained masked for the treatment allocation until the study database was locked. The study was conducted in accordance with the guidelines for Good Clinical Practice and the principles of the Declaration of Helsinki and was reviewed and approved by the Ethics Committee of Xinhua Hospital affiliated to Shanghai Jiao Tong University School of Medicine (XHEC-C-2016-103). Parents or legal guardians of all participants provided written informed consent. The trial was registered with ClinicalTrials.gov (NCT03156153).

### Outcome measures

The primary outcome was the change from baseline to 3-month in CARS (the total score ranges from 15 to 60 by summing up the 15 items, with a higher score indicating more severe autism)^[18]^. Confirmatory secondary outcomes were the change from baseline to 3-month in ADOS, SRS and the Clinical Global Impression-Improvement (CGI-I) to rate how much the patient’s illness has improved or worsened relative to a baseline measurement (a seven-point scale; 1 = “very much improved” to 7 = “very much worse”). SRS is applicable to children over 4 years old and therefore only a subset of children completed the SRS measures. The exploratory outcomes were the changes from baseline to 3-month in neurotransmitter (e.g. GABA and glutamate) concentrations within the IC. The safety outcomes were symptoms and blood parameters (serum potassium, uric acid, and creatine) closely monitored during the treatment period.

### Statistical analysis

Participants who received at least one dose of the study medication were included in the modified intention-to-treat population of primary outcome analysis and safety analysis, which was performed when the last trial participant reached 3 months.

The treatment effect of bumetanide was assessed by the change of the total score of CARS from baseline to 3-month using a mixed model ^[19]^. If the treatment effect was significant on the total score of CARS, we further tested the effects on 6 CARS items separately as suggested by our previous open-label study ^[15]^. Adjustment for multiple comparisons was performed at p = 0.05 using the false discovery rate (FDR) approach (Benjamin-Hochberg adjusted-p value). For CGI-I, the Kruskal–Wallis chi-squared test was applied to assess the significance level of the inter-group difference. Similarly, the treatment effects of bumetanide on the neurotransmitters in the insular cortex were tested by mixed models adjusted for age, sex, and intelligence. A permutation-based linear model was used to study the association between the changes of the neurotransmitter concentrations and the change of the CARS total scores, conducted by treatment group, adjusted for age, sex, intelligence, baseline CARS scores and neurotransmitter concentrations. If a significant association was detected, associations between the change in MRS measurement and the CARS items which showed significant treatment effects were further investigated and FDR correction was applied to control for multiple comparisons with items.

## Results

### Participants

From May 24th, 2017 through March 25th, 2019, of one hundred seventy-seven patients who were screened for eligibility, 120 were enrolled and 60 (50%) were randomly assigned to the bumetanide group and 60 (50%) to the placebo group (Figure 1). Among these 120 patients, one patient withdrew consent before the initiation of trial treatment and did not take any dose of study medication, resulting in 59 patients in the bumetanide group and 60 in the placebo group in the modified intend-to-treat analysis. Fifty-eight (97%) patients in the bumetanide group and 58 (97%) in the placebo group completed the 3-month study. One patient in the bumetanide group (who suffered from hand, foot, and mouth disease at week 4) and two in the placebo group (who were given the access to behavioural therapy at week 5) discontinued the treatment before the end of trial. For the MRS data, in the bumetanide group, 45 patients were scanned before the treatment, 38 after and 37 at both times due to MRI compliance and tolerance in young children. In the placebo group there were 49 patients scanned before the treatment, 43 after and 43 at both times. After the quality control of the MRS data, as described in the Methods section, the final sample consisted of 18 in the bumetanide group and 23 in the placebo group (Figure 1). The mean age of all patients was 4.12 years; 84% were male. The mean CARS total score was 38.17 (SD 4.08); 71% of all patients had severe symptoms with a total score exceeding 36 and a rating of 3 or higher on at least 5 of the 15 CARS items. Baseline characteristics such as age, sex, severity of ASD and IQ/DQ were well-balanced across the two groups (Table 1).

**Table 1:** Baseline characteristics of study participants

| **Table 1: Baseline characteristics of study participants** |  |  |  |  |  |
| --- | --- | --- | --- | --- | --- |
| *Variable* | *Bumetanide* | *Placebo* | *Degree of freedom* | *Statistic* | *p value* |
| N | 59 | 60 |  |  |  |
| Age, years | 4.03 (0.81) | 4.22 (0.94) | 115.0 | 1.17 | 0.25 |
| Sex |  |  |  |  |  |
| Female | 8 (14%) | 11 (18%) | 1 | 0.51 | 0.48 |
| Male | 51 (86%) | 49 (82%) |  |  |  |
| ADOS* |  |  |  |  |  |
| Communication | 6.41 (1.53) | 6.17 (1.65) | 116.6 | -0.82 | 0.41 |
| Social score | 10.83 (1.96) | 10.47 (2.22) | 115.6 | -0.95 | 0.34 |
| Social + Communication | 17.24 (3.28) | 16.63 (3.40) | 116.9 | -0.99 | 0.33 |
| Imagination | 2.75 (1.29) | 2.33 (1.24) | 116.6 | -1.77 | 0.08 |
| Repetition | 2.32 (1.44) | 1.92 (1.33) | 115.9 | -1.59 | 0.11 |
| ADOS (+) * | 59 (100%) | 60 (100%) |  |  |  |
| ADIR* |  |  |  |  |  |
| Total A | 21.22 (5.39) | 20.68 (6.12) | 115.6 | -0.51 | 0.61 |
| Total B non-verbal | 10.79 (3.25) | 10.40 (2.76) | 31.0 | -1.46 | 0.15 |
| Total B verbal | 12.59 (4.61) | 10.50 (3.79) | 79.9 | -0.58 | 0.56 |
| Total C | 6.22 (2.38) | 6.23 (2.25) | 116.4 | 0.03 | 0.98 |
| Total D | 3.22 (1.60) | 3.53 (1.67) | 116.9 | 1.04 | 0.30 |
| ADIR (+) * | 48 (82%) | 49 (82%) | 1 | 0.002 | 0.97 |
| CARS* |  |  |  |  |  |
| Total score | 38.27 (3.39) | 38.08 (4.68) | 107.6 | -0.26 | 0.79 |
| Number of items scored<br>≥3 | 6.20 (2.91) | 6.00 (3.97) | 108.1 | -0.32 | 0.75 |
| Severity of ASD* |  |  |  |  |  |
| Severe | 46 (78%) | 39 (65%) | 1 | 2.45 | 0.12 |
| Mild to moderate | 13 (22%) | 21 (35%) |  |  |  |
| Comorbidity with DD/ID* |  |  |  |  |  |
| With DD/ID | 44 (75%) | 39 (65%) | 1 | 1.29 | 0.26 |
| Without DD/ID | 15 (25%) | 21 (35%) |  |  |  |
| Data are mean (SD) or n (%). Group differences are evaluated with Welch's t test for continuous variables and Pearson's chi-squared test for categorical variables. *ADOS=The Autism Diagnostic Observation Schedule. *ADIR=The Autism Diagnostic Interview–Revised. *CARS=Childhood Autism Rating Scale. *Data shown in the ADOS (+) or ADIR (+) are the number and proportion of participants that met the criteria for ADOS or ADIR. *Severity of ASD was defined according to CARS: Severe symptoms of ASD were participants whose total score exceeded 36 and who had a rating of 3 or higher on at least 5 of the 15 items of CARS. *DD/ID=Developmental delay/intellectual disability: By intellectual assessment, the patients with developmental quotient (DQ) < 75 assessed using Gesell development scales, and intelligence quotient (IQ) < 70 assessed using WISC-R or WPPSI (Wechsler Intelligence Scale for children) were diagnosed as developmental delay or intellectual disability (DD/ID). |  |  |  |  |  |

**Figure 1:**
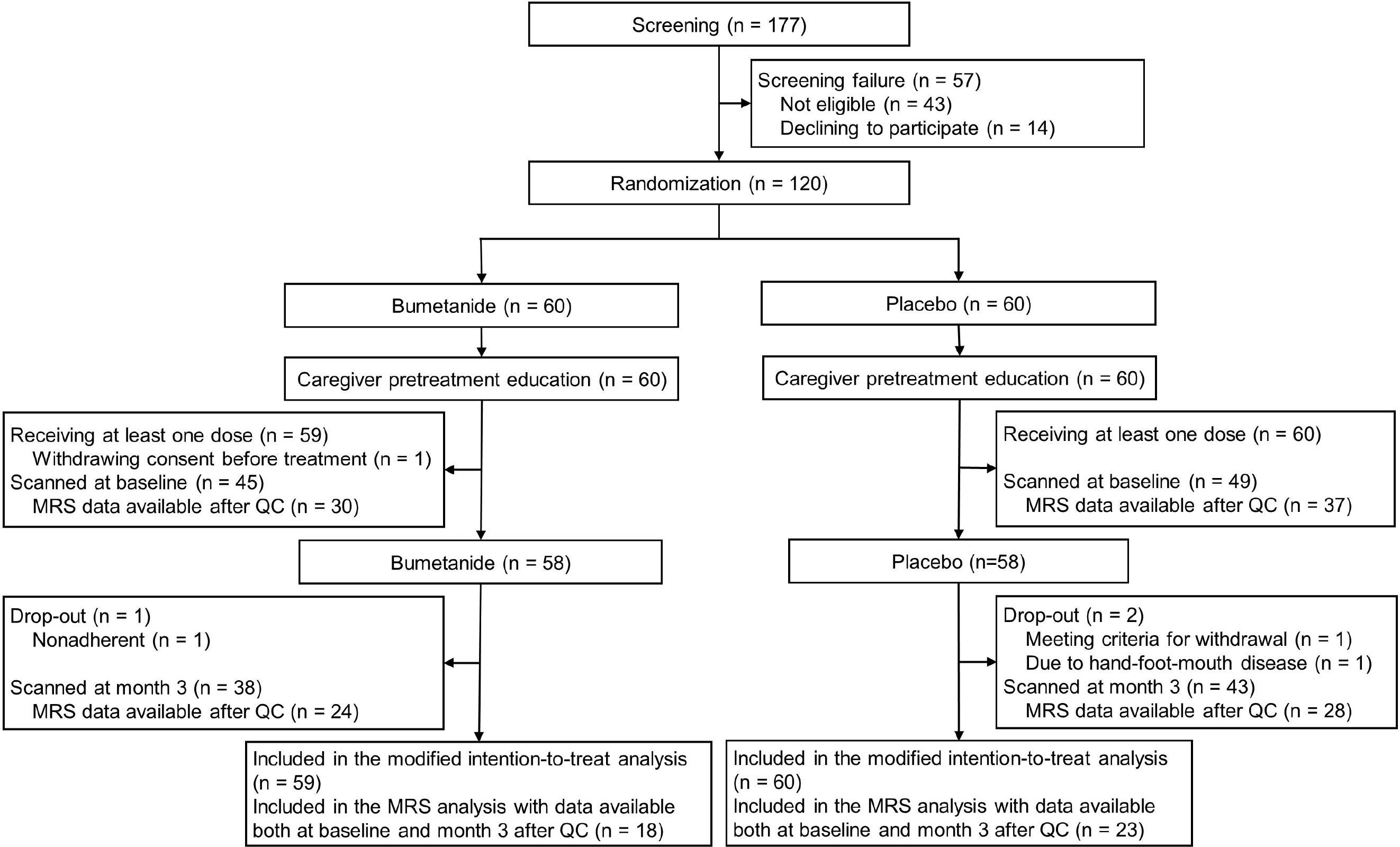
Trial profile. QC: Quality Control.

### Outcomes

In the modified intention-to-treat analysis, the total score of CARS decreased on average by 2.23 (SD 1.29) points in the bumetanide group and by 1.28 (SD 0.91) points in the placebo group. Using the mixed-effect model, the therapeutic effect of bumetanide compared with placebo was statistically significant (t_117_ = −4.67, p = 8.20×10^−6^; Figure 2a-b) and had a moderate-to-large effect size as measured by the partial-eta squared (η_p_^2^ = 0.15, Cohen’s D = 0.86 using a t-test of the group difference in the reduction of CARS total score). Significant decreases were also observed in the CARS items, including item 1 (impairment in human relationships) and item 13 (activity level); after adjustment for multiple comparisons item 1 (p-perm = 0.013, p-FDR = 0.039) and item 13 (p-perm = 0.011, p-FDR = 0.039) remained significant (Table 2). Compared with the placebo group, more patients in the bumetanide group had greater reductions of CARS total score and fewer patients had smaller reductions. The significance level of this difference between the distributions of the reduction of CARS total score after treatment was confirmed by the Kolmogorov–Smirnov test (D = 0.3, p = 0.0112; Figure 2c). Furthermore, this statistical difference in the reduction of CARS total score after treatment was supported by the analysis of the CGI-data for responders versus non-responders indicated with a trend (χ2 = 3.5, p = 0.061, supplementary Table 1). Moreover, a significant amelioration (t = −2.107, p = 0.0398, p-perm = 0.046) in SRS was shown in bumetanide group (11.16 (18.30)) comparing with that in placebo group (1.33 (16.27)), which provided an independent support for primary measure CARS (supplementary Table 5). However, no significant difference in ADOS was observed when comparing before and after 3-months treatment (supplementary Table 4).

**Table 2.**
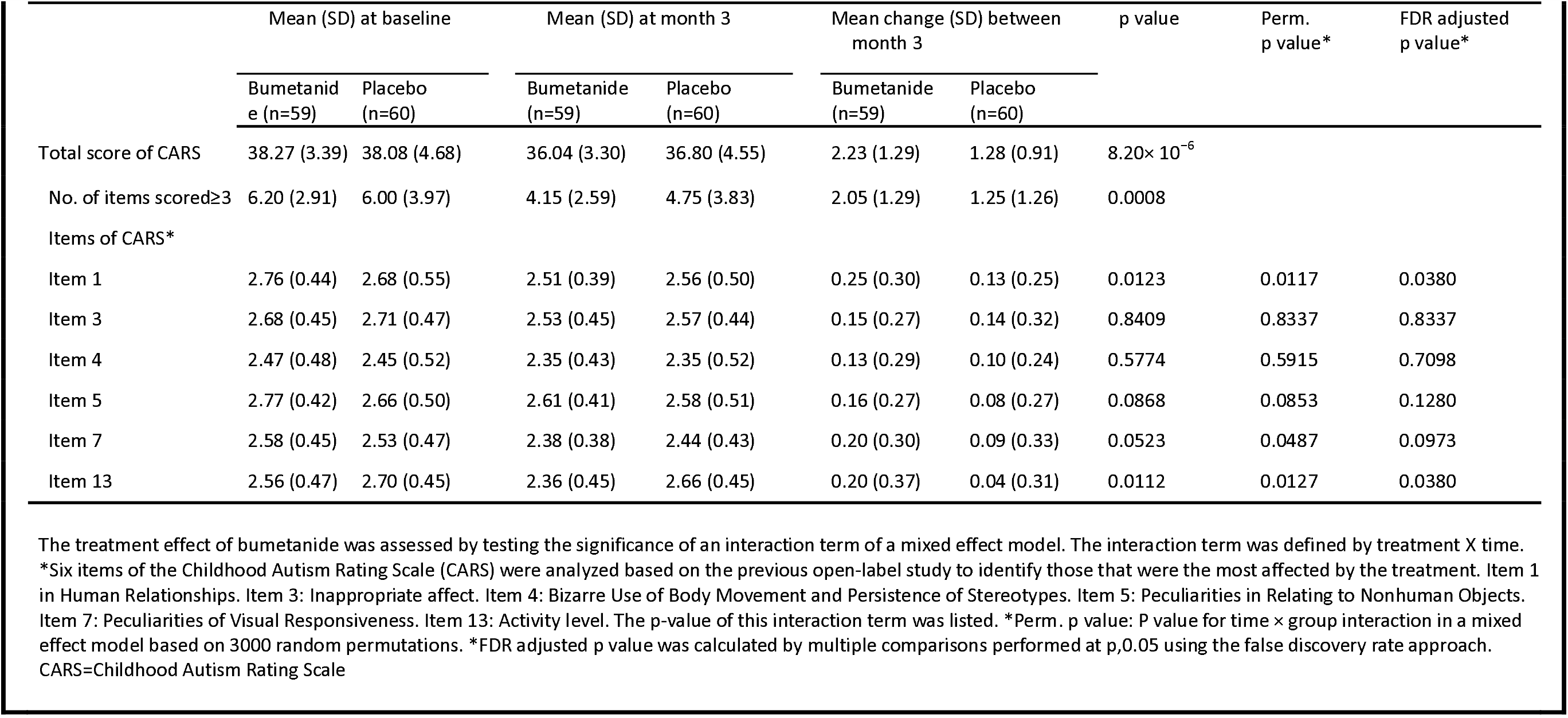
Primary clinical efficacy outcomes at month 3

**Figure 2:**
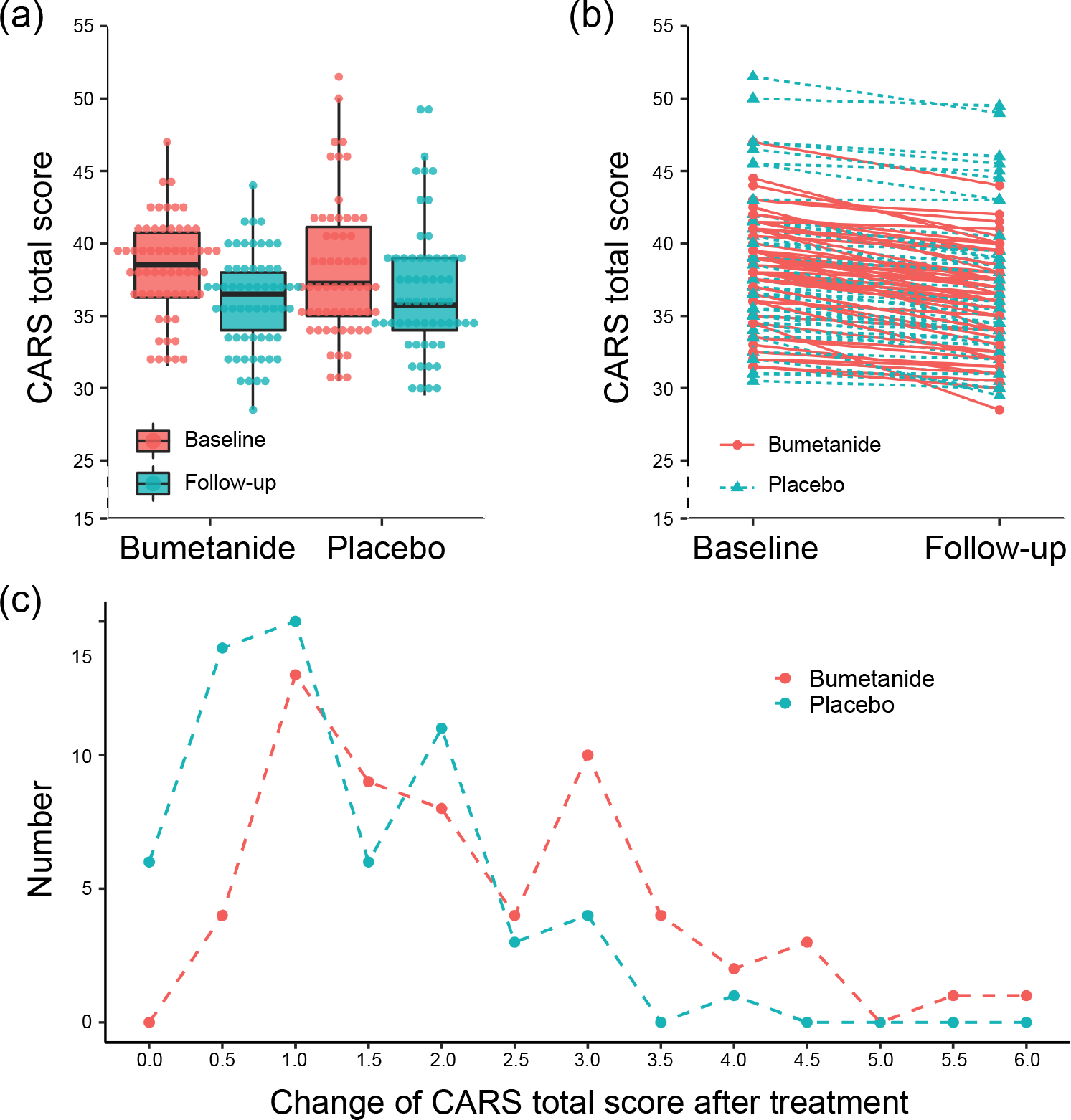
The change in CARS total score from baseline to 3-month follow-up. (a) Box plot and (b) line plot show the change in CARS total score from baseline to 3-month follow-up; (c) A line graph of the histogram values of the reduction of CARS total score after 3-month treatment for the bumetanide (in red) and the placebo (in green) groups.

The mean decrease between baseline and 3-month follow-up in the GABA/N-acetylaspartate (GABA/NAA) ratio in the IC was 0.0439 (SD 0.06) in the bumetanide group and 0.0002 (SD 0.06) in the placebo group. We found a significant difference in the changes of the GABA/NAA ratio between the drug group and the placebo group while controlling for the age, sex and IQ/DQ (t_36_ = −2.335, p = 0.025). Using the mixed-effect model, we found that bumetanide compared with placebo had a significant effect over the 3-month treatment course on the GABA/NAA ratio (t_40_ = −2.20, p = 0.034; p-perm = 0.04, η_p_^2^ = 0.08; Cohen’s D = 0.71 using a t-test of the group difference in the reduction of GABA/NAA ratio) (Figure 3a-b). Using the mixed-effect model, we also examined the NAA/Creatine (NAA/Cr) ratio and GABA/Cr ratio ^[20]^, and found no drug effect on NAA/Cr but a drug effect on GABA/Cr (t_38_ = −2.02, p = 0.051, p-perm = 0.04,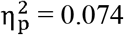)(supplementary Table 6). These results excluded the confounding effect of possible changes in NAA and further confirmed the drug effect on GABA. At the baseline before treatment, no significant difference in neither the GABA/Cr ratio nor the NAA/Cr ratio between the bumetanide group and the placebo group. The GABA change was predictive of the change in symptom severity as measured by the CARS total score (t_11_ = 2.278, p = 0.035) in the bumetanide group, but not in the placebo group (t_16_ = 0.483, p = 0.594) after the treatment (Figure 3c). For the specific symptom dimensions, the GABA change was predictive of the change in the activity level (t_11_ = 2.553, p = 0.009, p-FDR = 0.017).

**Figure 3:**
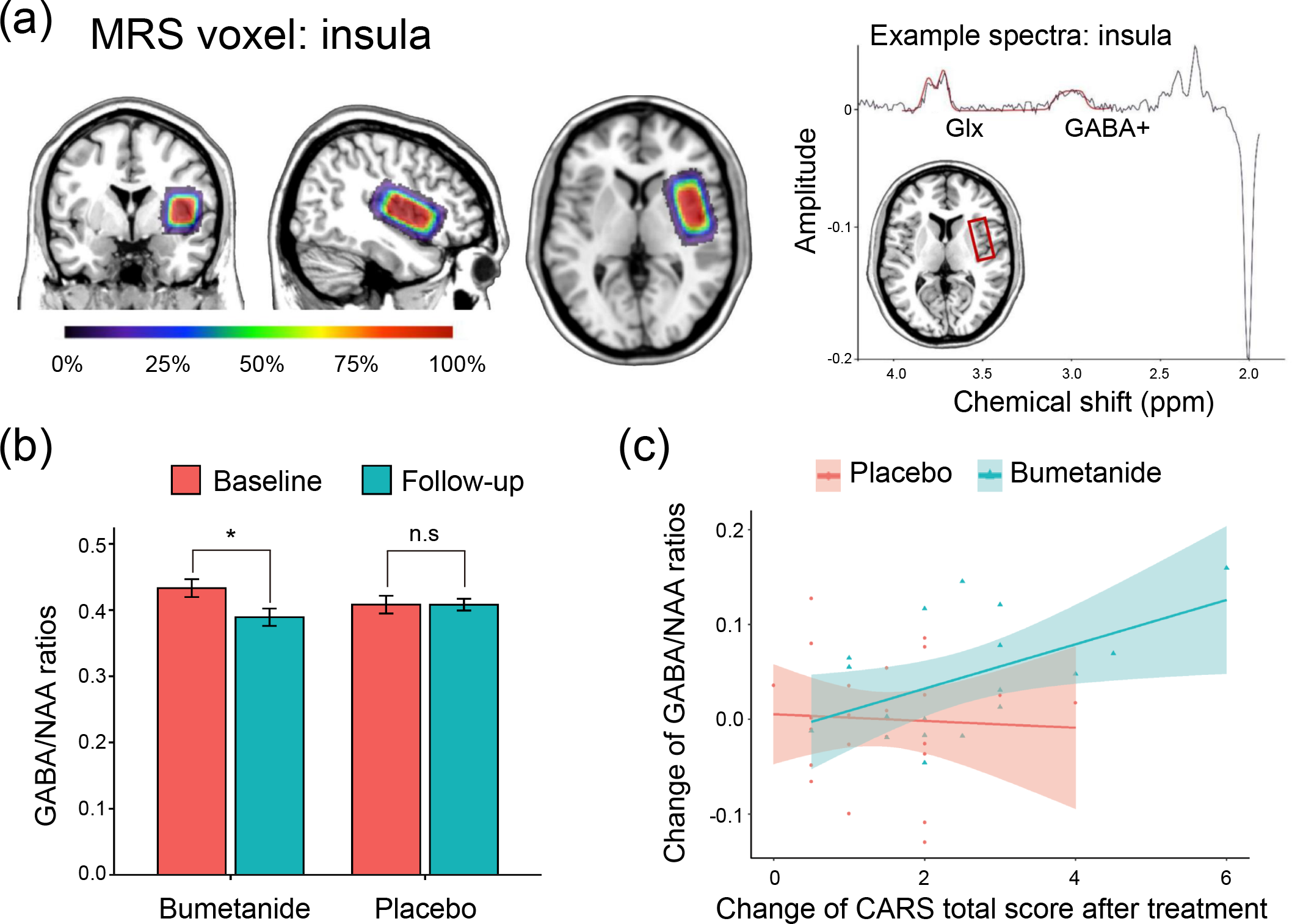
Changes in neurotransmitter levels after bumetanide administration. (a) Insular cortex VOIs in MRS. ppm: parts-per-million. (b) Box plot shows the changes in GABA/NAA ratios from baseline to 3-month follow-up in placebo and bumetanide groups.(c) The association of changes of GABA/NAA ratio with the changes of CARS total score after treatment for 3-month follow-up.

### Adverse events

All adverse events were graded 4 (mild) and no patient withdrew from the trial due to adverse events (Table 3). The most common adverse event observed was polyuria: 40 (67.8%) in the bumetanide group and 5 (8.3%) in the placebo group. However, there was no significant difference (t_57_=-0.29, p=0.77) between the decreases of CARS total scores in the subjects of bumetanide group with polyuria 2.26 (1.31) versus without polyuria 2.16 (1.28) (supplementary Table 7). There were 11 (18.6%) at 1-week, 4 (6.8%) at 1-month and 2 (3.4%) at 3-month in the bumetanide group developed mild hyperuricemia, and it was resolved by appropriate diet adjustment and more water drinking. A total of 6 in the bumetanide group (1 (1.7%) at 1-week and 5 (8.5%) at 3-month) developed mild hypokalemia, with serum potassium between 3.0 and 3.5 mmol/L. Hypokalemia was resolved after patients were administered potassium supplements and were advised to eat potassium-rich foods. The remaining side effects were loss of appetite (4 (6.8%) versus 1 (1.7%)), constipation (5 (8.5%) versus 2 (3.3%)), nausea (5 (8.5%) versus 2 (3.3%)), vomiting (1 (1.7%) versus 2 (3.3%)), diarrhea (0 versus 1 (1.7%)) and sleeping problem (1 (1.7%) versus 0) in the bumetanide group versus the placebo group (Table 3). Neither the change in blood potassium nor change in blood uric acid levels were associated with the decreases of CARS total scores (supplementary Table 8 & 9).

**Table 3.** Reported adverse events (in the modified intention-to-treat population)

|  | Bumetanide<br>(N=59) | Placebo<br>(N=60) |
| --- | --- | --- |
| Non-serious adverse event |  |  |
| Polyuria* | 40 (67.8%) | 5 (8.3%) |
| Loss of appetite | 4 (6.8%) | 1 (1.7%) |
| Constipation | 5 (8.5%) | 2 (3.3%) |
| Nausea | 1 (1.7%) | 0 (0%) |
| Vomiting | 1 (1.7%) | 2 (3.3%) |
| diarrhea | 0 (0%) | 1 (1.7%) |
| Sleeping problem | 1 (1.7%) | 0 (0%) |
| Mild hypokalemia* |  |  |
| 1-week | 1 (1.7%) | 0 (0%) |
| 1-month | 0 (0%) | 0 (0%) |
| 3-month | 5 (8.5%) | 0 (0%) |
| Mild hyperuricemia* |  |  |
| 1-week | 11 (18.6%) | 1 (1.7%) |
| 1-month | 4 (6.8%) | 0 (0%) |
| 3-month | 2 (3.4%) | 1 (1.7%) |
| Serious adverse events |  |  |
| Total | 0 (0%) | 0 (0%) |

Data are n (%). No participants withdrew after adverse events. \*Polyuria usually occurred within 3 hours of bumetanide treatment, which was resolved at the end of the treatment. \*Mild hypokalemia: Serum potassium usually between 3.0 and 3.5mmol/L. Mild hypokalemia was resolved after patients were administered potassium supplements and were advised to eat potassium-rich foods. \*Mild hyperuricemia usually occurred in one week or one month after the onset of treatment and was resolved by appropriate diet adjustment and more water drinking.

## Discussion and conclusion

In this randomised, double-blind, placebo-controlled trial of bumetanide in a large group (n = 120) of children with ASD aged 3 to 6 years, we found that bumetanide at a dose of 0.5 mg twice daily, significantly improved the core symptoms of ASD as measured by the primary clinical outcome (CARS total score), with no clinically significant side effects. There was a moderate to large effect size (η_p_^2^ = 0.15) of bumetanide treatment. We also found that the treatment of bumetanide reduced the insular GABA, and greater reduction in GABA was associated with greater decrease in the CARS total score in the patients who received bumetanide.

Although a number of early intensive behavioural interventions have been developed, the generalizability and availability remain a significant barrier for children with ASD after diagnosis ^[4]^. For example, in some European countries, more than 20% of children aged 7 years or younger receive no intervention at all.^6^ In China, the age of diagnosis is often between 3 and 6 years old, which is also a critical time window of rapid and dramatic postnatal brain development, and of fundamental acquisition of social and cognitive development^[21]^. The focus on children with ASD at an early age indeed may have the highest potential to improve social cognition and social learning deficits. However, there are fewer published trials in preschool-aged children with ASD than in school-aged children or adults. The children in the present trial were all aged 3 to 6 years, and the majority had severe ASD (71%) as determined by CARS total score. We conclude that the bumetanide is safe and effective in improving disease severity and attenuating social impairment and improving activity level for children, aged 3 to 6 years old, with ASD, and therefore provides a viable alternative therapeutic option for young patients in addition to behavioural interventions.

The therapeutic effects of bumetanide on autistic young children are encouraging, which calls for further rigorous studies on the underlying mechanisms. Based on the results, we suggest that the decreased GABA in the IC after bumetanide treatment in children with ASD mediates the improvement of core symptoms. The bumetanide-mediated reduction of insular GABA in the present study was positively associated with the decrease of CARS total score after the treatment with bumetanide. No significant treatment effect of bumetanide was observed on the glutamate concentration in the bumetanide group compared with the placebo group (t_39_ = −1.50, p = 0.143; Supplementary Method). This finding supports our previous open-label trial in 83 young patients which provided evidence that the symptom improvement following bumetanide treatment was significantly associated with reduced GABA in IC but not in visual cortex ^[15]^. We also found that the association of the insular GABA decrease was especially related to an improvement of a sensorimotor-related CARS item (item13 activity level), in keeping with our previous findings ^[15]^. According to DSM-5, the sensorimotor-related symptoms belong to the ASD core symptoms, and have been shown to precede and be predictive of social-communication deficits in childhood ^[22]^.

Bumetanide has been suggested to act on its molecular target Na-K-Cl cotransporter 1 (NKCC1) in the adult brain shown by animal studies ^[23, 24]^. NKCC1 acts as an inhibitor to reduce the intracellular chloride concentration ([Cl−]i) to promote the inhibitory nature of GABAergic synaptic transmission at the cellular level. Bumetanide has also been reported to hyperpolarize GABA reversal potential in immature neurons in mouse pups treated with intraperitoneal injections of bumetanide^[25]^. We found that bumetanide reduced insular GABA but not glutamate in autistic children, suggesting that bumetanide-mediated decrease in cortical GABA level might be considered as facilitating the restoration of GABAergic functioning, which is highly related to autistic core symptoms^[15]^. In addition, bumetanide as a NKCC1 inhibitor has been identified to preferentially acts on the GABAergic inhibitory interneurons over the glutamatergic excitatory cortical neurons in laboratory animals^[26]^, providing a potential mechanism for the specificity of GABA levels.

As a single-site investigator-initiated trial, the current study still has some limitations, including a relatively short treatment period (3 months) for bumetanide, which precluded an evaluation of long-lasting effect due to the short follow-up time. Thus, future studies with longer treatment periods would be helpful to evaluate the long-term therapeutic effects of bumetanide in children with ASD. In addition, although a pretreatment education session was given to all the participants to prevent any potential side-effect and ensure the implementation of blindness, some laboratory value changes (serum potassium and uric acid) may increase the risk of unblinding. However, the changes in laboratory values did not correlate with treatment response, which seems unlikely to have affected the results. Another minor limitation includes the lack of blinding validation, such as assessing whether the parents/caregivers knew whether the ASD child was on bumetanide or placebo. Consistent with previous studies, the present trial indicated that possibly not all young children with ASD will benefit from bumetanide treatment in such a short period (3-months). Due to the high heterogeneity in ASD, further investigations will be required to identify specific biomarkers for the potential subpopulation of effective responders to bumetanide treatment. It is worth mentioning that the CARS is used as a diagnostic tool. The clinical significance of change in CARS total score remains to be replicated in future studies utilising additional clinically sensitive measures to verify the clinical efficacy of bumetanide in children with ASD. In our MRS experiment, NAA was the only available peak acquired simultaneously to our metabolites of interest, namely GABA. To control for potential drifts in the spectra during acquisition and variations between scans, a simultaneously acquired reference peak is highly preferable. Therefore, we chose to compare the GABA/NAA ratio before and after treatment to assess the drug effect on GABA. It is theoretically possible that the drug effect detected for the GABA/NAA ratio was driven by NAA. However, the nonsignificant drug effect on both (Glx = Glutamate + Glutamine)/NAA and NAA/Cr but significant effect on GABA/Cr might suggest that the detected drug effect was unlikely to be driven by NAA. Future studies using more direct measurement of GABA and in additional brain regions may reveal more detailed effect of bumetanide in the autistic brains.

In summary, this trial in a large group of children with ASD aged 3 to 6 years old demonstrated that 3-month treatment of bumetanide at a dose of 0.5 mg twice daily is safe and effective in improving the core symptoms of ASD. In addition, we found that the therapeutic effect of bumetanide on disease was associated with changes in GABA in IC, which may help to elucidate the therapeutic mechanism of bumetanide.

## Data Availability

To facilitate reproducibility of our analysis, we have provided all data analytic code on GitHub in the following webpage: https://github.com/qluo2018/RCT.

## Acknowledgments

This project was funded by the Shanghai Municipal Commission of Health and Family Planning (No.2017ZZ02026, No. 2018BR33, No.2017EKHWYX-02 and No.GDEK201709), Shanghai Shenkang Hospital Development Center (No.16CR2025B), Shanghai Municipal Education Commission (No.20152234), National Natural Science Foundation of China (No. 81571031, No. 81761128035, No.81930095, No. 81873909, No.81701334, No. 81703249, No.82001771 and No. 31860306), Shanghai Committee of Science and Technology (No.17XD1403200, No.17ZR1444400, No.19410713500, No.18DZ2313505), Xinhua Hospital of Shanghai Jiao Tong University School of Medicine (2018YJRC03, Talent introduction-014 and Top talent-201603), National Human Genetic Resources Sharing Service Platform (2005DKA21300), the National Key Research and Development Program of China (2018YFC0910503), 111 Project (No.B18015), the Shanghai Municipal Science and Technology Major Project (No.2018SHZDZX01), Guangdong Key Project in “Development of new tools for diagnosis and treatment of Autism” (2018B030335001), Science and Technology Department of Yunnan Province (202001AV070010) and ZJ Lab.

## Evaluators

Mingyu Xu, MD PhD, Yun Fan, BA, Danping Wu, MS, Yongjun Chen, MS

## TSC

Kun Sun, MD PhD, Zhongchen Luo, PhD, Jiong Li, MD PhD, Ti-Fei Yuan, PhD, Li Yang, PhD

## DSMB

Lixiao Shen, MD PhD, Feng Li, MD PhD, Yongguo Yu, MD PhD, Wei Zhou, MD PhD, Xi Zhang, PhD

We also thank the children and families who participated in this study.

## Author contributions

Fei Li and Qiang Luo designed the study. Yuan Dai, Lingli Zhang, Yiting Ji, Kai Wang, Xujian Du, Yun Tang, Xin Liu, Hua He and Shining Deng collected the data. Yuan Dai, Lingli Zhang, Juehua Yu, Wei-Guang Li and Christelle Langley interpreted the data and wrote the report. Barbara J. Sahakian contributed to the data analysis and interpretation and manuscript preparation. Xin Zhou, Jun Zhang and Jianfeng Feng provided methodology support. Fei Li and Qiang Luo were the principal investigators. All authors approved the final version of the manuscript. Xinhua Hospital Shanghai Jiao Tong University School of Medicine, China was leading the study design and conducted the clinical trial; Fudan University, China was leading the biostatistics and MRS data analysis; University of Cambridge, UK team provided supports in study design, data interpreting strategy and article editing. Qiang Luo did the statistical analysis and signed it off. The statistical analysis plan was finalized on August 15, 2016. Data lock occurred on August 20, 2019. Xinhua Hospital Clinical Research Unit (CRU) provided financial administration. On-site audit was regularly carried out by the Shanghai Shenkang Hospital Development Center with no critical findings.

