## Supplemental Methods for "Improved symptoms following bumetanide treatment in children aged 3 to 6 years with autism spectrum disorder: a randomized, double-blind, placebo-controlled trial"

**APPENDIX**

**Appendix: Material & Methods**

The Material & Methods section comprises (i) the detailed method of the manuscript, (ii) the Rational to study 6 subscales of CARS, (iii) Power calculation of the sample size, (iv) Analysis of CGI-I interpreted as responder/non-responder, (v) Change of ADOS from baseline to 3-month follow-up, (vi) Change of RBS-R from baseline to 3-month follow-up, (vii) Training programs prior to MRS scans, (viii) Strategies to control the possible effects of chloral hydrate on MRS measurements, (ix) Testing the treatment effect of bumetanide on the glutamate concentration, as well as (x) MRI findings were not affected by a biased sampling due to the image quality.

***The detailed method of the manuscript***

Trial design and participant

The study used a randomised, double-blind, placebo-controlled, parallel-group design. We recruited eligible children from outpatient clinics at the Xinhua Hospital affiliated to Shanghai Jiao Tong University School of Medicine. Participants were considered eligible if they were diagnosed with ASD according to the Diagnostic and Statistical Manual of Mental Disorders, Fifth Edition (DSM-5); aged between 3 to 6 years; confirmed diagnosis with the Autism Diagnostic Interview–Revised (ADIR) and/or Autism Diagnostic Observation Schedule (ADOS), a CARS total score of no less than 30 and had no access to any behavioural intervention. We excluded patients if they had liver or kidney dysfunction; a history of allergy to sulfa drugs; abnormal electrocardiogram; genetic or chromosomal abnormalities; were diagnosed with neurological disease (e.g. epilepsy, Rett syndrome), or psychiatric disorder (e.g. very early-onset schizophrenia) other than ASD; severe hearing or visual impairment; were currently using melatonin for the treatment of sleep disorders or cessation of such treatment for less than 3 weeks. Additional exclusion criteria for neuroimaging were any contraindications of magnetic resonance imaging (MRI) scanning and any previous reports of traumatic brain injury. In regard to intellectual functioning, intelligence quotients (IQ), or developmental quotients (DQ) was assessed using the Wechsler Intelligence Scale for Children-Revised (WISC-R, Chinese version) for subjects ≥ 6 years old and the Wechsler Preschool and Primary Scale of Intelligence (WPPSI, Chinese version) for subjects < 6 but ≥ 4 years old, and the Gesell Developmental Schedules Chinese version for subjects < 48 months old.

The study was conducted in accordance with the guidelines for Good Clinical Practice and the principles of the Declaration of Helsinki and was reviewed and approved by the Ethics Committee of Xinhua Hospital affiliated to Shanghai Jiao Tong University School of Medicine (XHEC-C-2016-103). Parents or legal guardians of all participants provided written informed consent. The trial was registered with ClinicalTrials.gov (NCT03156153).

### Randomisation and masking

Participants were randomly assigned in a 1:1 ratio to receive bumetanide or placebo by use of a block randomisation scheme, with a block size of six. The generation of random allocation sequence and the preparation of trial medication were done by investigators in an external consultancy who do not participate in other aspects of the study. The study medication (bumetanide or placebo tablet) was provided in sequentially numbered envelopes. Bumetanide and placebo tablets were identical in appearance, smell and taste. Patients and their caregivers, investigators, experienced psychiatrists, and data analysts remained masked for the treatment allocation until the study database was locked. To reduce the possibility of differences in the incidence of polyuria between the two groups (bumetanide and placebo) potentially compromising the blinding effectiveness, a pediatrician was involved in the treatment of polyuria, but not in any other aspect of the study. This meant that the psychiatrist conducting the clinical ratings remained blind throughout the study.

### Intervention

Participants were given 0.5 mg oral bumetanide or placebo tablets twice daily for 3 months. Before that, a pretreatment education session of potential side effects of bumetanide (polyuria, hypokalemia, and hyperuricemia) and common symptoms (thirsty, fatigue, and loss of appetite) was delivered to the caregivers of the participants in both bumetanide and control groups. In addition, to minimize the impact of diuretic actions of bumetanide in blinding, a dietary supplement plan (adequate daily drinking water and potassium-rich diet) was encouraged for the applicants of both groups.

Outcome measures

The primary outcome was the change from baseline to 3-month in CARS (the total score ranges from 15 to 60 by summing up the 15 subscales items, with a higher score indicating more severe autism)(18). Confirmatory secondary outcomes were the change from baseline to 3-month in ADOS, SRS and the Clinical Global Impression-Improvement (CGI-I) to rate how much the patient's illness has improved or worsened relative to a baseline measurement (a seven-point scale; 1 = "very much improved" to 7 = "very much worse"). The exploratory outcomes were the changes from baseline to 3-month in neurotransmitter (e.g. GABA and glutamate) concentrations within the IC. Additional secondary and exploratory outcome measures included Symbolic Play Test, Chinese Communicative Development Inventory, and Short Sensory Profile Report scores as secondary outcomes, and electroencephalogram (EEG) recording, plasma metabolites, and GWAS as exploratory outcomes. These additional outcome measures will be reported in a subsequent study.

Adverse effect monitoring

Participants who received at least one dose of the study medication or placebo were assessed for safety. Symptoms (thirst, diuresis, nausea, vomiting, diarrhea, constipation, rash, palpitation, headache, dizziness, shortness of breath, and any other self-reported symptoms) were telephone interviewed at 1 week and 1 month. Blood parameters (serum potassium, uric acid, and creatine) were monitored via laboratory tests at 1 week and 1 month after the initiation of treatment and at the end of the treatment period. Adverse events that occurred on or after initiation of the study medication or placebo or preexisting medical condition that worsened during the treatment period were assessed and graded according to the World Health Organization standard.

MRI acquisition

Participants were scanned using a Siemens Verio 3.0-Tesla MRI scanner (Siemens Medical Solutions, Munich, Germany) with 32-channel head coil and four-channel neck coil. Scans were performed at chloral-hydrate-induced-sleep (19, 20). The dosage of chloral hydrate was 50 mg/kg up to a maximum dose of 1g administered rectally. Earplugs, earphones, and extra foam padding were provided to the subjects to reduce the sound of the scanner during the scan. The potential confounding effects of chloral hydrate were controlled for both by experimental design and analyses used.

The insula voxel (20 × 40 × 20 mm, Figure 2A) was placed along the anterior–posterior direction of the insular cortex and covered the anterior and posterior limits of the insula (21). To ensure the consistency of volume-of-interest (VOI) positioning in the longitudinal experiments, we used the first-scanned VOI of each participant as a reference to locate the same VOI in the follow-up scan. For each participant, a three-plane localizer image was first acquired, followed by a high-resolution anatomical T1-weighted magnetization-prepared rapid gradient echo image (192 sagittal slices; voxels = 1 × 1 × 1 mm; repetition time [TR] = 2300 ms; echo time [TE] = 2.28 ms; inversion time = 1100 ms; flip angle = 8°, field of view = 192 × 192 × 192 mm) to guide the spectroscopic VOI. For GABA measurement, Mescher-Garwood point-resolved spectroscopy (MEGA-PRESS) scan (256 spectra for the insular cortex were acquired with on-/off-resonance frequency = 1.9/7.5 ppm using TR/TE = 1500/68 ms) was performed in the VOI(22). The difference spectrum was obtained by subtracting the edit-ON and -OFF spectra, yielding a spectrum for total GABA and glutamate.

Image processing

We first used the LCModel software (23, 24) with a simulated MEGA-PRESS basis set to fit the MRS data, and then determined the n-acetylaspartate (NAA), n-acetylaspartyl-Glx, GABA, glutamine, and glutathione concentrations using the difference spectra, where GABA was actually GABA plus macromolecules around 3ppm and the NAAs we used was actually NAA plus NAAG. To ensure an acceptable signal-to-noise ratio (SNR) for the MRS voxel, we excluded those participants with SNR ≤ 15, full-width at half-maximum ≥ 0.05 ppm, and Cramer–Rao lower bounds in the fitted spectrum equal to or higher than 20% for GABA from the neuroimaging analyses. Figure 2A shows representative MRS spectra of the target VOI analyzed with the LCmodel.

We focused on GABA in this study, since our previous open-label study of bumetanide for children with ASD identified that the most significant change after the treatment was the decrease in GABA. This decrease in GABA was also found to relate to symptom improvement (15) We used the NAA concentration as an internal reference for the GABA concentration (25). Due to the individual variation in the tissue composition of the VOI, we further performed a tissue correction for the GABA-edited MRS to adjust the GABA measurements. In this paper, the GABA concentration refers to the corrected GABA metabolite concentration, i.e. GABA/NAA ratio corrected for tissue composition. As NAA was the only available peak acquired simultaneously to GABA in the MEGA-PRESS scans, we chose to use the GABA/NAA ratio as a proxy to the GABA concentration. To further test its validity, we calculated a GABA/Creatine ratio by combining GABA/NAA from the difference spectra with the NAA/Creatine from the edit-off spectra using the LCModel (26). Using the Statistical Parametric Mapping (SPM12) software, we estimated both the grey matter volume fraction (frGM) and the white matter volume fraction (frWM) within the VOI using the T1-weighte images (27). Therefore, the corrected GABA metabolite concentration was calculated by Metabolite-raw / NAA-observed × (1/[frGM + 0.5 × frWM]).

Statistical analysis

To detect a between-group difference of two or more points on CARS total score after 3-months treatment (primary outcome), a change from baseline of 3.1 SD and an alpha of 0.05, we required 51 participants per group to attain a power of 90%. Accounting for an expected no more than 15% drop out during follow-up period, we sought to include 120 patients (group size 60). Fifty-nine (98%) patients in the bumetanide group and 60 (100%) in the placebo group received at least one dose of the study medication, all of whom were included in the modified intention-to-treat population of primary outcome analysis and safety analysis, which was performed when the last trial participant reached 3 months.

The treatment effect of bumetanide was assessed by the change of the total score of CARS from baseline to 3-month using a mixed model (28). In this model, we assumed individualized random intercepts, and tested the treatment effect by the interaction term, treatment (0, placebo; 1, bumetanide) × time (0, baseline before treatment; 1, 3 months after treatment). The normality of the model residuals was assessed with the Shapiro-Wilk normality test, and homogeneity of variance across groups was evaluated with Levene's test. If at least one of the two tests were significant, a permutation-based mixed-effects model was established by 3000 random permutations of the group label using the permlmer function in the R package "predictmeans" v.1.0.1. If the treatment effect was significant on the total score of CARS, we further tested the effects on 6 CARS items separately as suggested by our previous open-label study (15), including item 1 (Impairment in human relationships), item 3 (Inappropriate affect ), item 4 (Bizarre use of body movement and persistence of stereotypes), item 5 (Peculiarities in relating to nonhuman objects), item 7 (Peculiarities of visual responsiveness), item 13 (Activity level: abnormal apathy or hyperactivity)(15). Adjustment for multiple comparisons was performed at $p = 0.05$ using the false discovery rate approach (Benjamin-Hochberg adjusted-p value). For CGI-I, the Kruskal–Wallis chi-squared test was applied to assess the significance level of the inter-group difference. Similarly, the treatment effects of bumetanide on the neurotransmitters in the insular cortex were tested by mixed models adjusted for age, sex, and intelligent (1, DQ<75 or IQ<70; 0, otherwise). A permutation-based linear model was used to study the association between the changes of the neurotransmitter concentrations and the change of the CARS total scores, conducted by treatment group, adjusted for age, sex, intelligence, baseline CARS scores and neurotransmitter concentrations, using the lmp function in the R package. If a significant association was detected, associations between the change in MRS measurement and subscales of the CARS items which showed significant treatment effects were further investigated and FDR correction was applied to control for multiple comparisons with items.

All analyses were conducted with the use of R v.3.5.1. To facilitate reproducibility of our analysis, we have provided all data analytic code on GitHub in the following webpage: https://github.com/qluo2018/RCT.

***Rational to examining 6 subscales of CARS***

The primary goal of this RCT was to test whether bumetanide had any treatment effect on the overall severity of ASD. The secondary goal was to test which symptom dimensions of ASD were significantly affected by bumetanide. Given that an open-label RCT without placebo control often shows larger treatment effect compared with a double-blinded RCT with placebo control, we focused on those 6 subscales of CARS that were significantly improved following the treatment with bumetanide in our previous open-label trial."

***Power calculation of the sample size***

This study was powered to detect a 3.8-point (SD 3.1) reduction in CARS total scored after 3-month bumetanide treatment, based on our preliminary intervention trial on 15 ASD patients, compared with 1.8-point in placebo group based on a previous publication [1], assuming the same SD as treatment. Setting the type I error rate to 0.05, a sample of 51 participants per group was deemed necessary to attain a power of 90%. Accounting for an expected no more than 15% drop out during follow-up period, we sought to include 120 patients (group size 60).

***Analysis of CGI-I interpreted as responder/non-responder***

We performed chi-squared test using CGI-I to categorize subjects as responders/non-responders. With 22 responders and 37 non-responders in bumetanide group and 13 responders and 47 non-responders in placebo group, the result only indicated a trend (p=0.061) between two groups.

***Change of ADOS from baseline to 3-month follow-up***

The Autism Diagnostic Observation Schedule (ADOS), designed as a diagnostic tool for ASD, is a semi-structured, standardized observation instrument, including domains of reciprocal social interaction, communication, and repetitive and stereotyped behaviours. All domains with higher scores indicate greater severity. The ADOS was assessed at baseline and after 3-month treatment. To test the treatment effect on ADOS, we used a mixed-effects model similar to that described in the main text for CARS. However, we did not observe significant difference in ADOS comparing before and after 3-month treatment.

***Change of RBS-R from baseline to 3-month follow-up***

The RBS-R (Repetitive Behavior Scale-Revised) is a scale to assess patient's repetitive behavior severity based on parent report. The RBS-R quantifies repetitive behavior in six behavioral subcategories: stereotypy, self-injury, compulsive behavior, ritualistic behavior, sameness, and restricted behavior. The RBS-R was assessed at baseline and after 3-month treatment. To test the treatment effect on RBS-R, we used a mixed-effects model similar to that described in the main text for CARS. However, we did not observe significant difference in RBS-R comparing before and after 3-month treatment.

***Change of SRS from baseline to 3-month follow-up***

The SRS is a quantitative measure of autistic traits in 4–18 years old. The SRS was assessed at baseline and after 3-month treatment on participants above 4 years old (25 in bumetanide group and 30 in placebo group). To test the treatment effect on SRS, we used a mixed-effects model similar to that described in the main text for CARS.

***Training programs prior to MRS scans***

Home training program: Carers were asked to help acclimatise the ASD children to using ear plugs and sleeping on their backs. Home training started 2 weeks prior to the MRI visit.

On-site training: 1) children were shown around the MRI scanning room to familiarise themselves with the new environment and various MRI sounds for 2-3 hours, prior to the actual scans; 2) children were brought to the MRI scanning room, they sat/lay on MRI scanner for around 10 min to become acclimatised to the MRI scanner and the room temperature, before falling asleep.

***Strategies to control the possible effects of chloral hydrate on MRS measurements***

The possible effects of chloral hydrate were controlled for both by experimental design and the analyses used. In the experimental design, both the children in the placebo group and the drug group received chloral hydrate. Therefore, the chloral hydrate was administered at both time points to both groups. In addition, in the analysis of the data, we used a mixed effect model instead of repeated measures ANOVA. By using this approach, we were able to subtract out the “time-invariant” effect of chloral hydrate.

***Testing the treatment effect of bumetanide on the glutamate concentration***

We also tested the treatment effect of bumetanide on the glutamate concentration. This concentration was measured by the Glx / NAA × (1/[frGM + 0.5 × frWM]), where Glx = Glutamate + Glutamine, and NAA = NAA + NAAG. The treatment effect was tested by the same mixed effect model as described in the main text. The change in the glutamate concentration in the bumetanide group after the 3-months treatment did not significantly different from that in the placebo group ($t_{39} = -1.50$, $p = 0.143$).

***MRI findings were not affected by a biased sampling due to the image quality***

We compared the bumetanide and placebo groups in terms of signal to noise ratio and other quality control parameters. No significant differences were observed (supplementary Table 2). We also compared the clinical features between the subjects with the MRS images which met and did not meet the quality control criteria. Again, no significant differences were detected (supplementary Table 3). Therefore, the MRS findings were strictly quality controlled and unlikely to be affected by a biased sampling, due to the image quality.

***The estimation of GABA+/creatine ratios***

GABA levels were firstly quantified relative to NAA as described in Method. Then the ratio of NAA to creatine was determined by fitting the spectral peaks in the edit-off spectrum with LCModel software. A set of metabolite basis spectra specific to the PRESS sequence (TE=68 ms) was used, provided by the MR Spectroscopy Lab (http://purcell.healthsciences.purdue.edu/mrslab/basis_sets.html). The ratios of GABA/NAA and NAA/creatine were combined to yield a GABA/creatine ratio (GABA/Cr), as previous reports [2, 3]. Of the subjects who had qualified GABA/NAA measurement, one was dropped when estimating GABA/Cr due to the missing of edit-off spectrum at baseline.
