## Supplemental Table 1-8 for "Improved symptoms following bumetanide treatment in children aged 3 to 6 years with autism spectrum disorder: a randomized, double-blind, placebo-controlled trial"

**Appendix: Supplementary tables**

**supplementary Table 1.** Difference in CGI-I between the bumetanide group and the placebo group at month 3

| | Bumetanide (n=59) | Placebo (n=60) | Group Comparisons | |
|---|---|---|---|---|
| | | | Kruskal-Wallis chi-squared | p value |
| **CGI-I** | | | 3.50 | 0.061 |
| Responder | 22(37.3%) | 13(21.7%) | | |
| Non-responder | 37(62.7%) | 47(78.3%) | | |

Data are n (%). CGI-I=Clinical Global Impressions-Improvement.

**supplementary Table 2.** Characteristics of MRS quality control related parameters

| Parameter | Baseline | | Follow-up | | Statistic | | | | | |
|---|---|---|---|---|---|---|---|---|---|---|
| | Placebo | Bumetanide | Placebo | Bumetanide | Baseline placebo vs Baseline Bumetanide | | Follow-up placebo vs Follow-up Bumetanide | | Baseline vs Follow-up | |
| Num. of images enter MRS Q.C. [a] | 49 | 43 | 43 | 37 | t/χ | p | t/χ | p | t/χ | p |
| FWHM (ppm) | | | | | | | | | | |
| Before Q.C. | 0.040 (0.019) | 0.039 (0.015) | 0.045 (0.022) | 0.044 (0.021) | 0.190 | 0.849 | 0.316 | 0.753 | -1.726 | 0.086 |
| Num. Passed Q.C. | 39 | 34 | 29 | 27 | 0.004 | 0.951 | 0.290 | 0.590 | 1.994 | 0.158 |
| SNR | | | | | | | | | | |
| Before Q.C. | 21.898 (4.214) | 20.721 (6.482) | 20.767 (5.259) | 19.703 (5.093) | 1.045 | 0.299 | 0.916 | 0.362 | 1.325 | 0.187 |
| Num. Passed Q.C. | 45 | 36 | 35 | 29 | 1.433 | 0.231 | 0.113 | 0.737 | 2.092 | 0.149 |
| GABA SD% | | | | | | | | | | |
| Before Q.C. | 8.327 (1.345) | 9.674 (5.498) | 8.140 (0.941) | 8.351 (1.338) | -1.661 | 0.100 | -0.828 | 0.410 | 1.583 | 0.115 |
| Num. Passed Q.C. | 49 | 41 | 43 | 37 | / | / | / | / | / | / |
| Num. Passed Q.C. of above three | 37 | 30 | 28 | 24 | 0.382 | 0.537 | 0.001 | 0.981 | 1.229 | 0.268 |

a: at baseline, two subjects with MRS/MRI image scanned did not enter the MRS quality control steps, one of them was the subject who did not enter the mITT analysis, the other one with the MRS insula voxel placed on the opposite hemisphere. At follow-up stage, one subject with MRS/MRI image scanned didn't enter these QC steps, as the MRS voxel placed on the opposite hemisphere.

**supplementary Table 3.** Characteristics of Subjects passed and failed in MRS quality control steps

| Variable | Baseline | | | | Follow-up | | | |
|---|---|---|---|---|---|---|---|---|
| | Placebo | Bumetanide | t/χ | p | Placebo | Bumetanide | t/χ | p |
| N | 49 | 43[a] | | | 43 | 37[b] | | |
| Age in years | 4.03(0.87) | 4.01(0.90) | 0.132 | 0.896 | 3.93(0.78) | 3.92(0.90) | 0.064 | 0.949 |
| Sex | | | | | | | | |
| Female | 9 | 7 | 0.069 | 0.792 | 8 | 7 | 0.001 | 0.971 |
| Male | 40 | 36 | | | 35 | 30 | | |
| CARS Total score | 37.81(4.32) | 38.44(3.59) | 0.770 | 0.443 | 36.52(4.09) | 36.23(3.35) | 0.353 | 0.725 |
| Comorbidity with DD/ID | | | | | | | | |
| With DD/ID | 31 | 31 | 0.812 | 0.368 | 27 | 28 | 1.537 | 0.215 |
| Without DD/ID | 18 | 12 | | | 16 | 9 | | |

a: at baseline, two subjects with MRS/MRI image scanned did not enter the MRS quality control steps, one of them was the subject who did not enter the mITT analysis， the other one with the MRS insula voxel placed on the opposite hemisphere.

b: follow-up stage, one subjects with MRS/MRI image scanned didn’t enter as the MRS voxel placed on the opposite hemisphere.

**supplementary Table 4.** Secondary clinical efficacy outcomes at month 3

| | Mean (SD) at baseline | | Mean change (SD) between baseline and month 3 | | p value | Perm. p value* | FDR adjusted p value* |
|---|---|---|---|---|---|---|---|
| | Bumetanide (n=59) | Placebo (n=60) | Bumetanide (n=59) | Placebo (n=60) | | | |
| ADOS | | | | | | | |
| Communication | 6.41 (1.53) | 6.17 (1.65) | 0.46 (0.74) | 0.20 (0.70) | 0.0476 | 0.0473 | 0.2366 |
| Social score | 10.83 (1.96) | 10.47 (2.22) | 0.52 (0.76) | 0.57 (1.45) | 0.8406 | 0.8417 | 0.9140 |
| Social + Communication | 17.24 (3.28) | 16.63 (3.40) | 0.96 (1.11) | 0.77 (1.40) | 0.3853 | 0.3999 | 0.6664 |
| Imagination | 2.75 (1.29) | 2.33 (1.24) | 0.25 (0.72) | 0.27 (0.75) | 0.9230 | 0.9140 | 0.9140 |
| Repetition | 2.32 (1.44) | 1.92 (1.33) | 0.18 (0.86) | 0.02 (0.86) | 0.3194 | 0.3182 | 0.6664 |
| RBS-R | | | | | | | |
| Stereotypy | 3.39 (2.24) | 2.97 (2.49) | 0.75 (1.36) | 0.67 (1.45) | 0.7591 | 0.7574 | 0.7574 |
| Self-injury | 0.47 (0.86) | 0.97 (1.38) | 0.14 (0.57) | 0.42 (1.08) | 0.0789 | 0.0750 | 0.3732 |
| Compulsive behavior | 3.19 (2.13) | 3.33 (2.36) | 0.64 (1.30) | 1.05 (1.69) | 0.1450 | 0.1599 | 0.3732 |
| Ritualistic behavior | 3.78 (2.39) | 3.77 (2.35) | 0.66 (1.82) | 0.90 (1.71) | 0.4618 | 0.4808 | 0.6732 |
| Sameness | 3.19 (2.47) | 3.42 (2.55) | 1.05 (1.89) | 0.95 (1.52) | 0.7488 | 0.7474 | 0.7574 |
| Restricted behavior | 3.2 (2.22) | 2.98 (1.96) | 0.42 (1.54) | 0.82 (1.50) | 0.1620 | 0.1529 | 0.3732 |
| Total | 17.22 (7.99) | 17.43 (8.12) | 3.66 (4.90) | 4.80 (5.25) | 0.2238 | 0.2309 | 0.4041 |

The treatment effect of bumetanide was assessed by testing the significance of an interaction term of a mixed effect model. The interaction term was defined by treatment X time. The p-value of this interaction term was listed. *Perm. p value: P value for time × group interaction in a mixed effect model based on 3000 random permutations. ADOS= Autism Diagnostic Observation Schedule. RBS-R= Repetitive Behavior Scale-Revised.

**supplementary Table 5.** Secondary clinical efficacy outcome (SRS) at month 3

| | Mean (SD) at baseline | | Mean (SD) at month 3 | | Mean change (SD) between baseline and month 3 | | t value | p value | Perm. p value* |
|---|---|---|---|---|---|---|---|---|---|
| | Bumetanide (n=25) | Placebo (n=30) | Bumetanide (n=25) | Placebo (n=30) | Bumetanide (n=25) | Placebo (n=30) | | | |
| Total | 88.12 (24.28) | 85.27 (18.13) | 76.96 (22.06) | 83.93 (20.38) | 11.16 (18.30) | 1.33 (16.27) | -2.1073 | 0.0398 | 0.0460 |
| Subscale 1 | 10.64 (2.93) | 10.40 (3.12) | 9.64 (2.81) | 10.57 (3.50) | 1.00 (3.28) | -0.17 (3.38) | -1.2911 | 0.2023 | 0.2109 |
| Subscale 2 | 17.28 (4.13) | 16.87 (4.04) | 15.36 (4.44) | 16.90 (5.57) | 1.92 (4.46) | -0.03 (4.11) | -1.6889 | 0.0971 | 0.1010 |
| Subscale 3 | 30.48 (9.72) | 29.13 (7.56) | 27.16 (8.82) | 29.17 (7.93) | 3.32 (8.09) | -0.03 (7.23) | -1.6227 | 0.1106 | 0.1136 |
| Subscale 4 | 15.12 (4.77) | 14.23 (4.06) | 13.20 (4.66) | 13.60 (3.37) | 1.92 (4.11) | 0.63 (3.55) | -1.2459 | 0.2183 | 0.2256 |
| Subscale 5 | 14.60 (7.11) | 14.63 (4.73) | 11.60 (5.92) | 13.70 (4.96) | 3.00 (5.00) | 0.93 (5.50) | -1.4462 | 0.1540 | 0.1423 |

The treatment effect of bumetanide was assessed by testing the significance of an interaction term of a mixed effect model.
The interaction term was defined by treatment X time.
The p-value of this interaction term was listed.
*Perm. p value: P value for time × group interaction in a mixed effect model based on 3000 random permutations.

**supplementary Table 6.** The NAA/Cr and GABA/Cr ratios at baseline and month 3

| | Mean (SD) at baseline* | | Mean (SD) at month 3 | | Mean change (SD) between baseline and month 3 | | t value | p value | Perm. p value |
|---|---|---|---|---|---|---|---|---|---|
| | Bumetanide (n=18) | Placebo (n=22) | Bumetanide (n=18) | Placebo (n=22) | Bumetanide (n=18) | Placebo (n=22) | | | |
| NAA/Cr | 1.53 (0.10) | 1.51 (0.10) | 1.53 (0.13) | 1.52 (0.09) | 0.00 (0.11) | -0.01 (0.10) | -0.22 | 0.82 | 0.77 |
| GABA/Cr | 0.56 (0.09) | 0.52 (0.09) | 0.50 (0.09) | 0.52 (0.06) | 0.06 (0.09) | 0.00 (0.08) | -2.02 | 0.05 | 0.04 |

*One subject who had qualified GABA/NAA measurement was excluded, due to the missing of edit-off spectrum

The treatment effect of bumetanide was assessed by testing the significance of an interaction term of a mixed effect model. The interaction term was defined by treatment X time.

The p-value and perm. p value (3000 random permutations) of this interaction term was listed.

**supplementary Table 7.** Difference in change of CARS total score between patients with polyuria and without polyuria in bumetanide group

| | Mean change (SD) between baseline and month 3 | | $t_{57}$ | p value |
|---|---|---|---|---|
| | With Polyuria (n=40) | Without Polyuria (n=19) | | |
| CARS total scores | 2.26 (1.31) | 2.16 (1.28) | -0.29 | 0.77 |

Shown are mean (SD). Group differences are evaluated with Welch's t test.

| supplementary Table 8 Correlations between change in laboratory tests and CARS scores in two groups | | | | | | | | | | | | |
|---|---|---|---|---|---|---|---|---|---|---|---|---|
| | blood potassium | | | | | | blood uric acid | | | | | |
| | bumetanide (n=59) | | | placebo (n=60) | | | bumetanide (n=59) | | | placebo (n=60) | | |
| CARS | r | p value | t value | r | p value | t value | r | p value | t value | r | p value | t value |
| Total score | 0.2776 | 0.1018 | 1.2177 | 0.0943 | 1.0000 | 0.4178 | -0.0012 | 0.3254 | -0.6506 | 0.0017 | 0.2704 | 1.1853 |
| No. of items scored ≥3 | -0.1491 | 0.9608 | -0.6456 | 0.3467 | 0.8039 | 1.1255 | -0.0007 | 0.6667 | -0.4093 | 0.0025 | 0.4265 | 1.2705 |
| Item 1 | 0.0450 | 0.4082 | 0.8445 | -0.0497 | 0.4110 | -0.7944 | -0.0001 | 0.8039 | -0.2551 | -0.0001 | 0.8235 | -0.2289 |
| Item 3 | 0.0238 | 0.5915 | 0.4995 | 0.0664 | 0.5000 | 0.8429 | -0.0002 | 0.5915 | -0.4084 | -0.0004 | 0.5915 | -0.7837 |
| Item 4 | 0.0272 | 0.7647 | 0.5296 | -0.0115 | 0.8235 | -0.1927 | 0.0001 | 1.0000 | 0.1238 | 0.0002 | 0.9020 | 0.5893 |
| Item 5 | 0.0301 | 0.6429 | 0.6240 | -0.0877 | 0.2281 | -1.3144 | 0.0000 | 0.8627 | 0.0886 | 0.0000 | 0.7059 | -0.1007 |
| Item 7 | 0.0408 | 0.9020 | 0.7733 | -0.0349 | 0.9412 | -0.4334 | 0.0000 | 0.9608 | 0.0601 | 0.0002 | 1.0000 | 0.4749 |
| Item 13 | 0.0376 | 0.8039 | 0.5638 | 0.0387 | 0.9412 | 0.5061 | 0.0007 | 0.5542 | 1.3525 | 0.0003 | 0.3223 | 0.6136 |

Shown are Pearson's correlation coefficients between the change in laboratory tests after treatment and the change in CARS scores.

**supplementary Table 10.** The GABA/NAA ratio at baseline and month 3

| | Mean (SD) at baseline [a] | | F value | p value | Mean (SD) at baseline [b] | | F value | p value |
|---|---|---|---|---|---|---|---|---|
| | Bumetanide (n=30) | Placebo (n=37) | | | Bumetanide (n=18) | Placebo (n=23) | | |
| GABA/NAA | 0.42 (0.06) | 0.52 (0.09) | 0.12 | 0.73 | 0.43 (0.06) | 0.41 (0.06) | 1.77 | 0.19 |

[a] Difference of data from two groups available after baseline quality control were examined.
[b] Difference of data from two groups available after both baseline and follow-up quality control were examined.
